# Machine Learning Prediction of Stroke Occurrence: A Systematic Review

**DOI:** 10.1101/2024.03.28.24305014

**Authors:** Sermkiat Lolak, Chaiyawat Suppasilp, Napaphat Poprom, Tunlanut Sapankaew, Myat Su Yin, Ratchainant Thammasudjarit, Gareth J. McKay, John Attia, Ammarin Thakkinstian

## Abstract

**Background:** Machine learning is nowadays commonly used for disease prediction, including cardiovascular disease. There is growing evidence of the effectiveness of machine learning algorithms for stroke risk prediction models.

**Aims:** A systematic review was conducted to identify and comprehensively evaluate the available evidence.

**Summary of review:** Relevant studies were identified from the three electronic databases (i) MEDLINE via Pubmed, (ii) Scopus, and (iii) IEEE *Xplore* from inception to 1^st^ December 2020. Out of 12,626 studies identified, 40 used machine learning for ischemic or hemorrhagic stroke risk prediction models. Synthesis without meta-analysis identified that a boosting algorithm (median C-statistics = 0.9 (interquartile range [IQR]: 0.88-0.92)), and neural network (median C-statistic = 0.80 (IQR: 0.77-0.92)) performed best among ML models in the low risk of bias studies. Moreover, a boosting algorithm also performed best in overall (both low and high risk of bias) studies (median C-statistic = 0.92 (IQR: 0.90-0.95)).

**Conclusions:** The systematic review found promising results of the ML algorithm model performances compare with the gold standard conventional models, such as FSRP (C-statistic 0.653) and revised FSRP (C-statistic 0.716). In term of the algorithm, boosting and neural networks are robust, but are considered as black-box models, since they are composed of non-linearity and complex algorithms. It remains questionable whether a physician would adapt these algorithms to use in a real clinical setting. Moreover, less than half of the studies (16 out of 40) were at low risk of bias in our systematic review. More researches with good methodology and study design, alongside explainable and good performance models, may become available in the future.

**Trial Registration Information:** The International Prospective Register of Systematic Reviews (PROSPERO) database (ID: CRD42021234081).

## Introduction

Stroke is commonly characterized by a sudden focal injury to the central nervous system, primarily a cerebral vascular event.^1^ Apart from significant stroke-associated mortality, morbidity can vary from minimal deficit to severe, including the bedridden or vegetative states. Seven million adults were reported having a stroke in the United States between 2013 and 2016,^2^ approximately 795,000 people experiencing new episodes or recurrent strokes per year equating an incidence of 2.5% mostly (87%) due to ischemic stroke.^3^ Stroke represents a significant healthcare burden given the significant morbidity, and also its impact and suffering to relatives.^4^ Stroke prevention can significantly reduce this burden. For effective prevention, it is necessary to accurately assess individual stroke risks.

There are multiple risk factors associated with stroke, given the different pathological pathways involved in the disease process.^5^ While the Framingham Stroke Risk Profile is a widely used model which predicts the 10-year likelihood of stroke risk for men and women based on eight risk factors (i.e., age, systolic blood pressure, antihypertensive therapy, diabetes mellitus, cigarette smoking, cardiovascular disease, atrial fibrillation, and left ventricular hypertrophy),^6^ the INTERSTROKE group suggested that these modifiable risk factors were responsible for 90% of the stroke population-attribute risk.^7^ Foremost, hypertension is the most important modifiable risk factor of hemorrhagic stroke, whereas smoking, diabetes mellitus, hypercholesterolemia, and cardiac concerns are more critical factors in ischemic stroke. Globally, high systolic blood pressure is a leading single risk factor for developing stroke (57%; 95% CI: 49.8% - 64.4%), while metabolic risks (e.g., body mass index, systolic blood pressure, plasma glucose, cholesterol level) accounted for 72.1% (95% CI: 66.4% - 77.3%).^8^

Some of the risk factors of stroke are not modifiable (e.g., ethnicity, age, sex, etcetera), whereas others are modifiable (e.g., hypertension, diabetes mellitus, smoking, dyslipidemia, alcohol consumption, etcetera), which could be improvable thereby enabling improved risk factor control. Several risk prediction models have been developed from general adult populations (e.g., Framingham Stroke Risk Profile (FSRP),^6^ revised FSRP,^9^ or Copenhagen City Heart Study,^10^ and from specific disease (e.g., HAS-BLED,^11^ CHAD_2_ score,^12^ CHAD_2_S_2_-VASc,^13^ etcetera for atrial fibrillation). Most of the widespread risk prediction scores were developed using conventional statistical approaches using regression modeling, which assumes linear associations, but that may not always be a valid assumption.^14^ In addition, those risk factors may modify each other, and so might limit conventional regression approaches. In addition, the conventional model is sometimes limited to appropriate capture of non-linear relationships. That is one concern which can be addressed through machine learning (ML) approaches.^15^ Nevertheless, ML algorithms do not always offer improved modeling beyond traditional statistical approaches.^16,17^

Nowadays, there is advanced and widespread use of ML models in multiple healthcare fields. In addition, predictive modeling using MLs and data mining of real-world data (RWD) (e.g., electronic health record (EHR), health surveys, administrative claims data) is growing, as well as the development of stroke risk prediction modeling. However, there are many aspects to consider while developing and validating a risk prediction model (e.g., type of study population, study design, variables and measures, model algorithm, model evaluation, etcetera), which can affect the model performance and applicability but varied between studies. Though there are growing papers using ML in stroke risk prediction models, they are less widely applied in clinical practice. This might be due to multi-factors including research quality, the explainability of the ML model, different settings, and lack of external validation. Therefore, a systematic review of stroke risk prediction models developed from the ML models was conducted to identify the key features used, type of ML models, study phase (derived, internal/external validations), and model performance in ischemic and hemorrhagic strokes.

## Methods

The Preferred Reporting Information for Systematic Reviews and Meta-Analysis (PRISMA) protocol was followed to conduct this systematic review which was registered with the International Prospective Register of Systematic Reviews (PROSPERO) database (ID: CRD42021234081).

### Patients and Public Involvement

Patients and the public were not involved in any way.

### Search strategy

The author (S.L) was in charge of searching the relevant studies from three electronic databases (i) MEDLINE via Pubmed, (ii) Scopus, and (iii) IEEE *Xplore^®^* since inception to 1^st^ December 2020. Search terms and strategies were constructed based on the Patients, Intervention, Comparator, and Ooutcome framework^18^; see Supplemental Table 1.

### Study selection

The search results were imported to the EndNote^tm^ 20. The author (S.L) and another group of reviewers (C.S, N.P, T.S, M.Y) independently screened the titles and abstracts based on the eligibility criteria. Inconsistent results were discussed among the team, disagreements were resolved by a member of the second group not involved in the screening.

Studies published in any language were eligible if they met following criteria: Constructed a stroke risk prediction model in adults (≥18 years), had at least two risk features considered within the predictive model, applied any ML algorithm, reported model performance in terms of calibration or discrimination, and had the outcome as acute ischemic or hemorrhagic stroke.

Studies were excluded with the following criteria: aimed to assess the association between risk features and stroke without constructing a risk prediction model, had outcome as various stroke (e.g., venous sinus thrombosis, Moyamoya syndrome, or hematologic disease), combination of stroke with others (e.g., cardiovascular disease (CVD), major advanced cardiovascular event (MACE), acute myocardial infarction, or peripheral arterial disease), published in non-English language which could not be translated by Google translation.

### Data extraction and assessment of risk of bias

Two reviewers (S.L and C.S) independently extracted study design (e.g., pro-retro-spective cohorts, nested case-control), study phase (derived, internal/external validations), type of outcome and measures, follow-up time, predictors (e.g., numbers, types (continuous / categorized), timing, selection), missing data handling techniques, models (type of statistical or ML models), discrimination performance (C-statistics, sensitivity, specificity, precision, accuracy), calibration performance, and interpretation. (e.g., using the model compared to other studies).

### Quality assessment

The risk of bias (RoB) was assessed independently by two reviewers (S.L. ad C.S.) using the prediction model study risk of bias assessment tool (PROBAST) checklist.^19^ The tool evaluates four domains (i.e., participants, predictors, outcomes, and analysis). Each domain assesses both RoB and applicability, except the analysis domain that only assesses RoB. We graded the overall RoB and applicability as ‘low’ if all domains were rated as a low, ‘high concern’ if at least one of the domains was regarded as high, and ‘unclear’ if there was insufficient information. The inconsistent grades were solved by discuss and consensus between two reviewers.

### Data synthesis

Findings of reviews were described following the guideline of Synthesis without meta-analysis (SWiM).^20^ The model performance included C-statistics, sensitivity, specificity, and accuracy which were described by study phases and RoB groups.

## Results

### Study search

A total of 12,626 records were identified, of which 40 studies met the eligibility criteria, and reasons for ineligibility are described in Figure 1. We obtained a high level of agreement between reviewers (97.6%).

**Figure 1.**
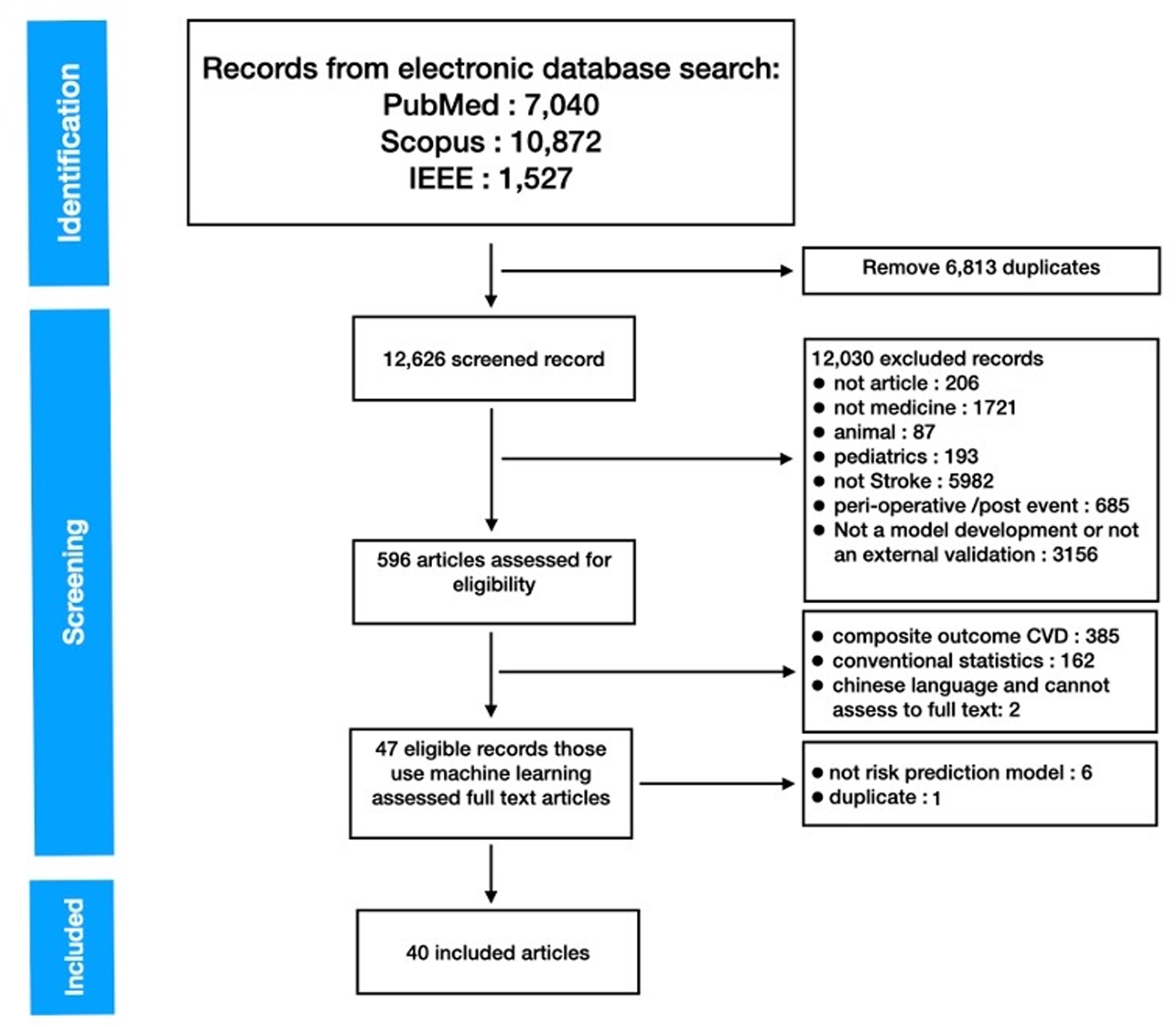
PRISMA Flow diagram.

### Dataset and methodology

Supplemental Table 2 summarizes the characteristics of the 40 studies included. A total of 2,340,341 patients were included from the 40 studies, with 26,929 stroke events. However, several studies used the same public dataset, e.g., the Kaggle data.^21–27^ The median number of included risk features was 14 (range: 6 - 2007), with a median follow-up time of three years. Most studies were based on model derivation phases, and only a single study performed both derivation and external validation phases. Most studies included participants from all age ranges, while four studies focused only on the elderly (age 65 or older) and a single study on younger participants (age 25-45 years). In terms of risk participants with underlying disease, two, three, one, and two studies included only carotid stenosis, atrial fibrillation, hypertension, and recurrent stroke, respectively. In terms of dataset sources, 16 and 16 studies were single-center and multicenter sources, and eight studies used data from Kaggle public ML datasets (https://www.kaggle.com/datasets) and University of California, Irvine (UCI) (https://archive.ics.uci.edu/ml/index.php). The largest dataset was from the National Health Insurance Research Database (NHIRD), Taiwan,^28–30^ which was electronic medical claimed data. Of the 40 studies included, the study designs were retrospective cohorts (N=24), prospective cohorts (N=4), case-controls (N=9, and nested-case-controls (N=3). All features used for developing models were structural data.

Regarding the missing data, 27 of the 40 studies did not either clearly report the extent of missing data nor how the missing values were dealt. Among the 13 studies that considered the missing data, seven used deletion, and three used mean imputation, whereas the rest studies used various methods of imputation. Less than half of the included studies had reported participant characteristics, see Supplemental Table 2.

### Risk of Bias

Details of the studies’ participants and designs for evaluating PROBAST domains are provided in Supplemental Table 3. Many studies fell in unclear RoBs for the analysis domain due to multiple criteria relating to this domain including described how the model was developed and validated, how the performance was measured, or how missing values were reported and handle, etcetera. Regarding the missing data issues, the most common method identified included deleting or using mean imputation given most studies did not clearly state how the missing data were treated. Therefore, 19 and 18 studies were of ‘low concern’ in the participant and analysis RoB domains, respectively, while nine studies were of ‘low concern’ for overall judgment of both RoB and applicability, see Figure 2. Inappropriate or missing follow-up timing made some studies fall into the high or unclear outcome RoB domain, while poor assessment and evaluation degraded some studies in predictor RoB domain. We also considered the relaxed criteria of low-RoB, if studies had no more than a single ‘unsure’ overall RoB applicability concern, and 16 studies fell within this category.

**Figure 2.**
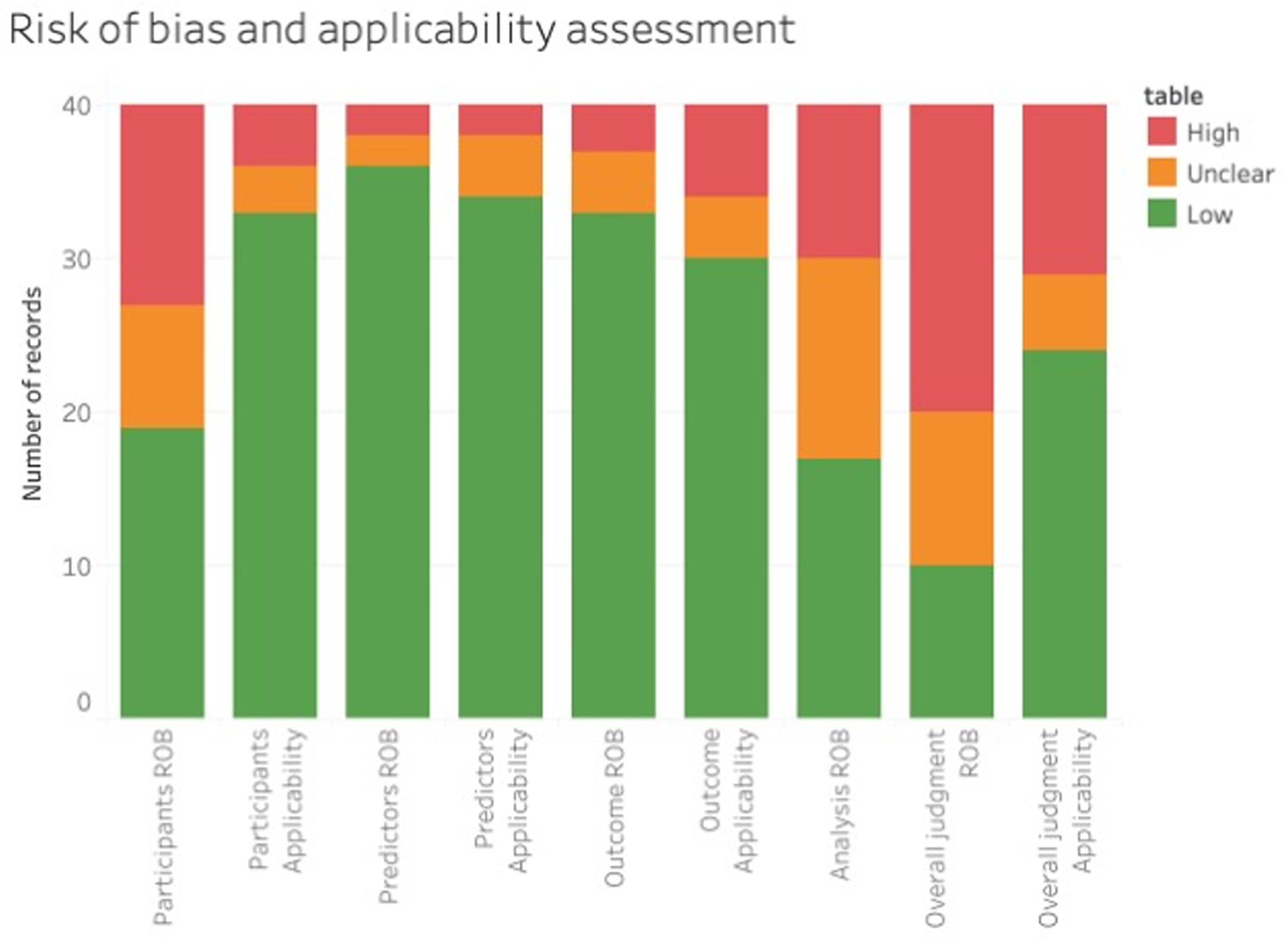
Summarized assessment for risk of bias and applicability by PROBAST domain.

Additionally, concerning the PROBAST evaluation, ten, ten, and twenty studies were categorized as low, unsure, and high overall judgment for RoB, respectively. There were twenty-four, five, and eleven studies in the low, unsure, and high RoB categories for applicability, see Figure 2 and Supplemental Table 4. Details of RoB stratified by model are provided in ML algorithms subsection below.

### Feature selection and feature engineering

Various methods were applied for feature selection, see Supplemental Table 5. Hence, the most common structural features used were age, gender, hypertension, diabetes mellitus, smoking, alcohol consumption, history of CVD, body mass index (BMI), and blood sugar. Four studies used forward/backward eliminations,^31–34^ six studies used principal component analysis (PCA),^23,24,35–38^ four studies used information gain,^28,38–40^ and three studies used the Chi-Square test to select the relevant risk factors or features^27,41,42^; and the rest used various methods, e.g., correlation, least absolute shrinkage and selection operator (LASSO), specific algorithm, and medical knowledge from standard guidelines, etcetera.

### ML algorithms

The seven most frequent models used were (i) Neural network (NN) in 24 studies (10 studies were low RoB) (ii) Support vector machine (SVM) in 20 studies (8 studies were low RoB), (iii) Decision tree (DT) in 12 studies (7 studies were RoB) (iv), Regression in 11 studies (7 studies were low RoB), (v) Random Forest (RF) in 10 studies (8 studies were low RoB), (vi) Naïve Bayes in 9 studies (3 studies were low RoB), and (vii) Boosting in 6 studies (5 studies were low RoB). Considering only for the best outcome, or the preferred model if there was no comparison, 15 studies used NNs, nine used SVM, six studies used only regression, and two studies each used RF, DT, and Boosting. NN architecture is usually comprised of 1-3 hidden layers with varying of the number of neurons.

### Evaluation methods

Most studies use standard data splitting for evaluation. Nineteen studies split the data into train-test or train-validation-test sets with ratios of 60:40 to 90:10. A total of 14 studies used 5- or 10-folds cross-validation, two used leave-one-out cross-validation, and the remaining did not report. In terms of evaluation metrics, most studies used common evaluation metrics for risk prediction model, with C-statistics or area under ROC curve, sensitivity, specificity, and accuracy; while fewer reported precision, F-score. ^21,22,26,27,32,34,36,39,41,42^

A summary of the evaluation metrics is described in Supplemental Table 5. The boosting algorithm provided the best overall median C-statistic of 0.92 (IQR: 0.90-0.95), followed by SVM [median C-statistic= 0.85 (IQR: 0.74-0.94)] and NN [median C-statistic= 0.78 (IQR: 0.75-0.91)]. In low-RoB studies, the boosting algorithm remained the best performing with a median C-statistic of 0.90 (IQR: 0.88-0.92), followed by NN with a median C-statistic of 0.80 (IQR: 0.77-0.92).

Concerning sensitivity, RF and Boosting both performed well in all studies with median values of 0.89 (IQR: 0.88-0.92) and 0.89 (IQR: 0.89-0.92), respectively, with both ML models performing well in the low RoB group, with corresponding median values of 0.89 (IQR: 0.88-0.92) and 0.89 (IQR: 0.87-0.90). For specificity, the SVM and logistic regression models performed best in all studies with median values of 0.82 (IQR: 0.75-0.98) and 0.79 (IQR: 0.70-0.89), respectively. The SVM and NN models performed best in the low RoB group with median values of 0.75 (IQR: 0.74-0.79) and 0.75 (IQR: 0.33-0.80). One study provided model calibrating with Hosmer-Lemeshow test.^39^ Summarized model performances are in Figure 3.

**Figure 3.**
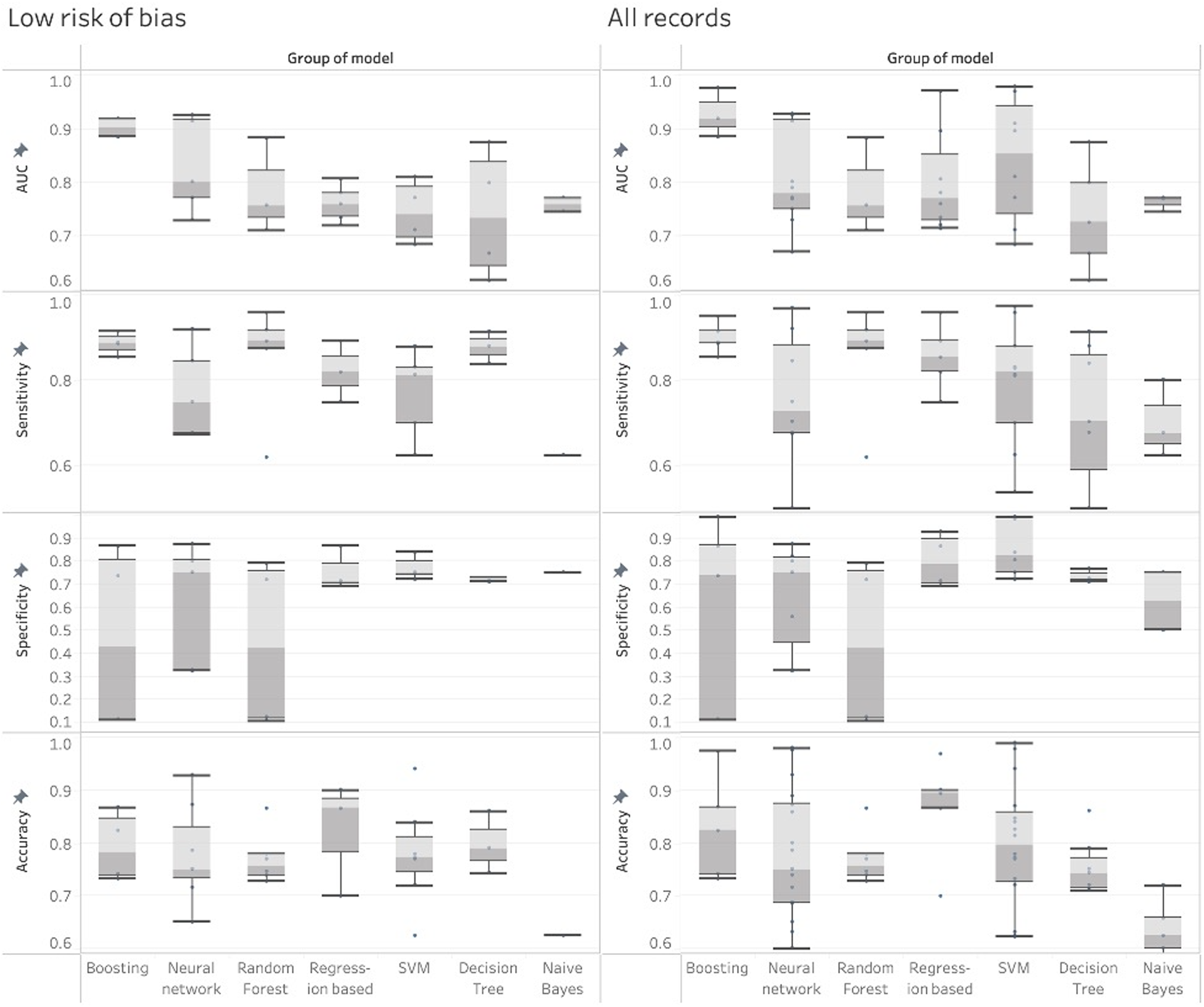
Box plots of evaluation metrics of overall included studies and studies with low risk of bias by type of models.

The key features vary between studies and the dataset. Most common key features are age, gender, smoking, heart disease, hypertension, BMI, diabetes mellitus, blood sugar, and work type. Details are provided in Supplemental Table 6.

## Discussion

A systematic review was performed to evaluate ML performance for stroke risk prediction modeling. NN, SVM, and decision tree are the three most common ML models, while logistic regression was used both in ML and conventional statistical models. The number of features ranged from 6 to 2007, while the most common data splitting is train-test splitting and 10 folds cross-validation, with one study external validating the model.

Comparing ML algorithms was complicated due to the heterogeneity of datasets, with different features and methodologies used. Despite the SWiM method can provide a direction for data synthesis,^20^ it still lacks a strong numerical procedure as a pulling method in a meta-analysis. Nevertheless, this systematic review gave some insights into the ML stroke risk prediction model. Our review found promising results of the ML algorithm performances compare with the gold standard conventional models, such as FSRP (AUC=0.653) and revised FSRP (AUC=0.716). Among ML algorithms, it was found that the boosting and NN algorithms performed best in terms of discriminative performance C-statistics. In a CVD meta-analysis, Krittanawong *et al*.,^43^ reported that SVM, boosting algorithm, and convolution neural network had the highest pooled C-statistic for stroke prediction. At the same time, this analysis yielded a close result between boosting and SVM in general studies. NNs, like most MLs, are versatile insofar as they can capture non-linear associations and are more robust than a standard statistical model to the effects of multicollinearity between included parameters, meaning the weighting of individual parameters is of less importance coupled with some loss of explainability.^44^ The gradient boosting tree model is also considered robust to the features correlated since the model grows a small tree that works greedily so that the redundant features are not selected. NNs are comparable to boosting, when there is sufficient data with good fine-tuning of the hyperparameters. The discriminatory C-statistics offers measures of model sensitivity and specificity, with the former particularly important. A false positive (patients that did not have a stroke, but were predicted by the model as having a stroke) is preferred to a false negative (patients did have a stroke, but were classified as not having a stroke by the ML model). When compared between studies, it was also necessary consider this issue. The sensitivity and specificity trade-off depend on the cut-off value, so it is reasonable to look at the C-statistics first. The higher the discriminatory power of C-statistics, the lower the sensitivity and specificity trade-off. In this review, RF is another ML algorithm that achieved good sensitivity. RF generates a ‘forest’ of decision trees with a random subset of features. However, overall performance was diminished by low specificity or reported high false-positive findings. There is a concern regarding the black-box nature of ML approaches, given the lack of understanding around the variables combined within the model. These findings indicate that the best-performing models (e.g., NN, SVM, RF, Boosting) were the models whose results cannot be explained easily. In contrast, regression analysis is considered a transparent parametric modeling approach, but with poorer performance. Further research is ongoing to explain ML and AI approaches better and improves model transparency. Lipton^45^ discussed the “understanding” and “interpretability” in many aspects as they might have paramount differences for the user concerning which part they try to understand. While Montavon^46^ focused on post-hoc interpretability and gave definitions of “interpretation” and “explanation”; *Interpretation* is mapping to human concepts which are able to understand, such as text or image’s pixels input are interpretable, but abstract features such as word embedding are not. On top of *interpretation*, an *explanation* is a relevance score given to each interpretable feature, such as a heatmap or features’ weight. Regarding this concept, a decision tree classifier is more explainable due to the solid scores given in hierarchical order, while NNs might need a heatmap which varies between individual data. In a clinical setting, physicians and patients need an explainable model, so they can focus on giving a certain treatment and make modifications to lifestyles to lower the risk of the disease. The standard risk prediction model, such as FSRP has a scoring of each feature, which is highly and directly explainable and still widely used. Two studies compared the ML model with FSRP, and the ML model gave a better performance in exchange for the loss of explainability. ^39,47^ However, recent research on model interpretation such as LIME, ^48^ SHAP, ^49^ or developing transparent models such as InterpretML, ^50^ or AIX360 ^51^ might help clarify and make the models more explainable. We still haven’t noticed many papers recognizing the importance of explainability in the ML risk prediction model.

Heterogeneity of data used for model development is one of the major cause of diverse results. Different data have different sets of features which resulted in different performance. For example, many studies used public datasets such as Kaggle, resulting in more weight inputting on their available features in overall key features pooling. We strongly encourage more external validation research to overcome this heterogeneity problem. On the other hand, most of the studies included cross-sectional data, which lacked time to event evaluation afforded by longitudinal data. Time to the event is important given the significance of age as a major risk factor. Age and length of follow-up period represent important timing features, especially for individuals attending regular health check-ups, particularly at a young age or early-stage disease onset. The inclusion of such information would likely improve model performance. Only fourteen studies were found that reported the follow-up time, see Supplemental Table 2. Chen-Ying *et al*., ^28^ use timestamp of features for temporal representation, i.e., 6-month or 1-year value. Although the time can be integrated into the model, feature correlation and multicollinearity will occur. Feature selection and choice of ML algorithm can help combat a correlation problem as we described above, but again, explainability might be lost. Moreover, the influence of many risk factors varies over time, and consideration of such changes within dynamic ML modeling would offer a more accurate reflection of real-world data related to individual-level stroke risk. It should be encouraged with modeling approaches, where sufficient data is available. Longitudinal data modeling using a specific model such as Linear Mixed Model or Recurrent Neural Network (RNN) could be beneficial.

Many of the studies included also had imbalanced datasets, which reflected the real-world situation that only minor patients develop stroke in the dataset. It might be hard for the model to learn an effective function to predict stroke regarding this issue. Ten studies used up or down-sampling techniques, ^21,23,24,26,27,29,30,37,52,53^ such as a synthetic minority over-sampling technique (SMOTE) to balance or smooth the data, ^54^ while others did not. Care is required when evaluating highly unbalanced data. The C-statistic and model accuracy may reflect a false level of competency if the positive class is a common class, but this is less likely in some clinical events. A better metric for future consideration would include the precision-recall (PR) curve in low incidence diseases,^55^ such as stroke, since the PR curve considers both positive in terms of recall and negative classes in terms of false-positive while varying its threshold. In other words, the PR curve switches from false-positive rate (FPR) in the ROC curve to precision, which is a different view since low FPR often means a better model, but low recall means a worse model. Since an imbalanced dataset tends to have a low FPR by its nature, using a ROC curve might provide an overoptimistic view of the model.

Like many systematic reviews, the quality of studies included was of concern. Many of those considered had high RoBs, with only 16 out of 40 studies within our research considered in the low-RoB group. Also, many studies failed to report any participant characteristics (Supplemental Table 3), with only 17 out of 40 providing information, though many only provided partial reporting. A further cause for concern from publicly available data is the lack of or limited information available regarding data collection, especially if unpublished. For instance, a high proportion of missing data for some variables, such as smoking status in the Kaggle dataset, where 30% was missing. In this instance, the lack of explanatory data obscures the mechanism of missingness and determination of whether this may be due to randomness or otherwise,^56^, especially where current smokers may be reluctant to answer, in contrast to missingness due to random events. Modeling under such circumstances and a lack of knowledge surrounding data attributes would likely lead to limited sensitivity and specificity. A large, transparent, and open public real-world dataset is needed to tackle this problem.

Feature selection and feature engineering form the center of ML modeling, although they contrast insofar as the former tends to reduce, while the latter tends to multiply. Feature engineering is an approach widely used in ML prediction which aims to change the angle of how the model looks at the data. Feature selection is an essential tool to limit issues associated with multicollinearity, especially with highly correlated risk features. That type of model can also prove influential, with NN less affected by multicollinearity, with regression modeling more vulnerable. Details of feature selection are presented in Supplemental Table 5. For instance, Li et al.,^57^ used various feature selection methods and ML algorithms to construct a two-year risk prediction model for ischemic stroke and other thromboembolism events in atrial fibrillation patients achieving a higher C-statistic (0.71-0.74) than the standard Framingham or CHA_2_DS_2_-VASc models (0.66-0.69) using the standard cox/logistic regression model. PCA is another popular approach with one of the included studies comparing *a priori* and *a posteriori* MedDietScore using PCA-derived food groupings through ML models, which generated comparable C-statistics for both approaches, although multiple logistic regression was only slightly improved.^35^

Even if NNs address the issue of multicollinearity, they are still limited by the lack of transparency or the rationale for feature inclusion, which is especially important in healthcare settings. Using the causal feature to avoid or limit potential confounders is preferential. Although more studies using NNs may offer improved clarity or explainability compared to the past, we were unaware of any that met the inclusion criteria for our systematic review. Sometimes, the predictive performance is prioritized by policymakers, with the most robust model accuracy preferred over feature explainability. Nevertheless, for health promotion or individual treatments, both clinical transparency and accuracy are still preferred to provide patient clarity concerning modifiable behaviors.

Model validation is crucial for developing risk prediction models to combat the overfitting problems, yet we found only a single study that had undertaken external validation and several studies that provided internal calibration. Data sharing should be encouraged to facilitate independent external model validation to test generalizability. More studies are expected in specific disease classes with higher stroke risk, such as atrial fibrillation, carotid stenosis, or transient ischemic attack. Furthermore, the novel biomarker, e.g., N-Terminal Pro-B-Type Natriuretic Peptide,^58^ C-reactive protein,^59^ could be considered in future studies where available as it is considered a standard recommendation for CVD screening. Real clinical adaptation in stroke prediction using ML models is still lacking since it is still not yet in the standard practice guideline. A randomized controlled trial, a web or mobile application, a large high quality, and diverse study as well as recognizing of technology in the medical community would help the physician to adapt the technology to use and conduct more studies in real clinical settings. Our findings should be scrutinized in light of several limitations. Firstly, the RoB in ML studies was high, especially concerning the participants’ domain. Many studies used public datasets with insufficient information on data collection or participant characteristics. Additionally, only eight out of 40 studies reported the accuracy in the analysis domain, which could mislead inexperienced readers, particularly for imbalanced datasets.

## Conclusion

This systematic review found that the boosting and NN algorithms can help develop robust stroke risk prediction models. Notwithstanding, both models lose some clarity through a lack of explainability of the features included as a tradeoff, which is an important healthcare issue in research and application. Nevertheless, research on this issue is growing, and we expect to see improved transparency and more robust modeling in the future. Data quality and quantity and the research methodological approaches are essential issues to generate more accurate risk prediction modeling, particularly in real-world clinical settings.

## Supporting information

PRISMA checklist

## Acknowledgment

This manuscript is part of Sermkiat Lolak’s course completion of Ph.D. in Data Science for Healthcare (International Program), Department of Clinical Epidemiology and Biostatistics, Faculty of Medicine Ramathibodi Hospital, Mahidol University. The authors kindly thank Mr. Stephen John Pinder and Nattakrit Tongpoonsakdi (QLS Certificate No. 1603177) for conducting a thorough English language review on our manuscript. We would like to extend our deepest gratitude to Prof. Peter Haddawy for his invaluable insights, constructive feedback, and unwavering support throughout the development of this manuscript. His expertise and mentorship have significantly contributed to the quality of this work, and we are truly appreciative of his dedication and guidance.

## Supplemental material

**Supplemental information** is available for this paper.

## Data availability

Data sharing is not applicable to this article as no new data were created or analyzed in this study.

## Author contributions

This study has been initiated and conceptualized by SL, RT, PH, and AT. Review methodology was designed by RT and AT. SL developed search terms and performed search. Also, SL screened all titles and abstracts as a first reviewer. CS, NP, TS, MY acted as the second group of reviewers for screening the articles. SL and CS independently extracted the data and assessed the risk of bias. The manuscript was drafted by S.L and revised by MY, GM, PH, JA, RT, and AT. All review processes were supervised by AT. All authors have approved the final version of this manuscript.

Correspondence and requests for materials should be addressed to Dr. Ratchainant Thammasudjarit.

## List of Supplemental Tables

- Supplemental Table 1: Search strategy
- Supplemental Table 2: Participants’ characteristics.
- Supplemental Table 3: Participants and study design.
- Supplemental Table 4: Assessment RoB of each study by PROBAST domain
- Supplemental Table 5: Evaluation metrics.

## Abbreviations

AI: Artificial Intelligence
BMI: Body Mass Index
CVD: Cardiovascular disease
DT: Decision tree
EHR: Electronic Health Record
FPR: False Positive Rate
FSRP: Framingham Stroke Risk Profile
IQR: Interquartile Range
LASSO: Least Absolute Shrinkage and Selection Operator
MACE: Major Advanced Cardiovascular Event
ML: Machine Learning
NHIRD: the National Health Insurance Research Database
NN: Neural network
PCA: Principal Component Analysis
PRISMA: Preferred Reporting Information for Systematic Reviews and Meta-Analysis
PROBAST: The Prediction model study Risk of Bias Assessment Tool
PROSPERO: The International Prospective Register of Systematic Reviews
RCT: Randomized Controlled Trial
RoB: Risk of Bias
RF: Random Forest
RWD: Real-world Data
SMOTE: Synthetic Minority Over-sampling Technique
SVM: Support vector machine
SWiM: Synthesis without meta-analysis
UCI: University of California, Irvine

