## Supplemental Table for "Machine Learning Prediction of Stroke Occurrence: A Systematic Review"

Tables

**Table 1: Search strategy**

| Search | Query | Results |
| --- | --- | --- |
| 1 | ("stroke"[MeSH Terms] OR "stroke"[Title/Abstract] OR "strokes"[Title/Abstract] OR "stroke's"[Title/Abstract]) | 295,035 |
| 2 | "intracerebral haemorrhage"[Title/Abstract] OR "cerebral hemorrhage"[MeSH Terms] OR ("cerebral"[Title/Abstract] AND "hemorrhage"[Title/Abstract]) OR "cerebral hemorrhage"[Title/Abstract] OR ("intracerebral"[Title/Abstract] AND "hemorrhage"[Title/Abstract]) OR "intracerebral hemorrhage"[Title/Abstract] | 58,437 |
| 3 | "ich"[Title/Abstract] | 11,599 |
| 4 | ((“predic*"[Title/Abstract] OR "progno*"[Title/Abstract]) AND "model*"[Title/Abstract]) OR ("risk index"[Title/Abstract]) OR ("clinical score"[Title/Abstract])) AND ("risk*"[Title/Abstract]) | 124,093 |
| 5 | ("machine learning"[Title/Abstract]) OR ("support vector machine"[Title/Abstract]) OR ("Support Vector Machine"[Title/Abstract]) OR ("SVM"[Title/Abstract]) OR ("neural network"[Title/Abstract]) OR ("Neural Network"[Title/Abstract]) OR ("Random Forest"[Title/Abstract]) OR ("random forest"[Title/Abstract]) OR ("Deep Learning"[Title/Abstract]) OR ("deep learning"[Title/Abstract]) OR (machine learning[MeSH Terms]) | 92,596 |
| 6 | #4 OR #5 | 212,808 |
| 7 | (#1 OR #2 OR #3) AND #6 | 7,040 |

**IEEE Xplore**

| Search | Query | Results |
| --- | --- | --- |
| 1 | (("All Metadata":intracerebral haemorrhage) OR "Mesh_Terms":cerebral hemorrhage OR  (("All Metadata":cerebral) AND ("All Metadata":hemorrhage)) OR "Search_All":cerebral hemorrhage OR (("All Metadata":intracerebral) AND ("All Metadata":hemorrhage)) OR "Search_All":intracerebral hemorrhage) | 148 |
| 2 | (("Mesh_Terms":stroke) OR "All Metadata":stroke) | 12,927 |
| 3 | ("All Metadata":ich) | 203 |
| 4 | (((("All Metadata":predic*) OR ("All Metadata":progno*)) AND ("All Metadata":model*)) OR ("All Metadata":risk index) OR ("All Metadata":clinical score)) AND ("All Metadata":risk*) | 11,528 |
| 5 | ("Mesh_Terms":machine learning) OR ("All Metadata":machine learning) OR ("All Metadata":SVM) OR ("All Metadata":support vector machine) OR ("All Metadata":random forest) OR ("All Metadata":neural network) OR ("All Metadata":deep learning) | 350,661 |
| 6 | #4 OR #5 | 359,439 |
| 7 | (#1 OR #2 OR #3) AND (#6) | 1,527 |

**SCOPUS**

| Search | Query | Results |
| --- | --- | --- |
| 1 | TITLE-ABS ( ( intracereb*  OR  cereb*  OR  brain )  AND  ( hemor*  OR  infarc*  OR  ischem* ) ) | 195,368 |
| 2 | TITLE-ABS ( stroke  OR  strokes  OR  stroke's  OR  cerebrovascular  OR  cva*  OR  ( brain  AND  vascular ) ) | 428,868 |
| 3 | TITLE-ABS ( ich ) | 25,554 |
| 4 | (((TITLE-ABS ( predic* ) OR TITLE-ABS ( progno* ))  AND  TITLE-ABS ( model* ) ) OR  TITLE-ABS ( “risk index” ) OR TITLE-ABS( “clinical score” )) AND TITLE-ABS(risk*) | 176,166 |
| 5 | TITLE-ABS ( "machine learning" )  OR  TITLE-ABS ( "support vector machine" )  OR  TITLE-ABS ( "svm" )  OR  TITLE-ABS ( "neural network" )  OR  TITLE-ABS ( "random forest" )  OR  TITLE-ABS ( "deep learning" ) | 736,062 |
| 6 | #4 OR #5 | 903,143 |
| 7 | (#1 OR #2 OR #3) AND (#6) | 10,872 |

**Table 2: Participants’ characteristics.**

| **Author (Year)** | **Mean Age** | **% Female** | **Mean BMI** | **% Current Smoking** | **% DM** | **% HT** | **Mean SBP** | **% Anti HT drug** |
| --- | --- | --- | --- | --- | --- | --- | --- | --- |
| Kyriacou *et al.* (2015) | 81 |  |  |  |  |  |  |  |
| Kastorini *et al*. (2013) | 77,  73 | 44.4, 44.4 | 26.72, 27.35 | 19.6, 18.9 | 32.9, 21.5 | 84.4, 56.8 |  |  |
| Hung *et al*. (2019) | 35.5 |  |  |  | 5.7 | 11.9 |  |  |
| Chan *et al*. (2019) | 71,  65 | 50, 43.8 |  | 20,  34.5 | 32.5, 29.9 | 72.5, 62.3 | 166, 160 | 47.5, 50.9 |
| Orfanoudaki *et al. (*2020) | 54.41 | 54 | 27.17 | 26 | 6 | 0.7 | 127.3 | 0.2 |
| Ayyagari *et al*. (2014) | 67.3, 58.2 | 52.1 |  |  | 41, 24.2 |  | 136.13,  134.95 | 28.8, 26.9 |
| Ray *et al*. (2020) |  | 72 |  |  |  |  |  |  |
| Wu *et al*. (2020) |  | 51.64 |  | 20.78 | 2.3 | 25.46 | 142 |  |
| Zhang *et al.* (2019) | 64.68 | 58.1 |  |  |  |  |  |  |
| Yonglai *et al*. (2017) |  | 59 |  |  |  |  |  |  |
| Colak *et al*. (2015) |  | 46.5 |  |  |  |  |  |  |
| Jeena *et al*. (2019) | 66 | 49.4 | 26 |  | 16.7 |  | 160 |  |
| Albu *et al*. (2019) | 47.08 | 47 |  |  |  |  |  |  |
| Clifford *et al*. (2019) | 70.3 | 47.2 |  | 20.3 | 25.7 | 73.5 |  |  |
| Arslan *et al*. (2016) | 53.97 | 47.4 |  |  |  |  |  |  |
| Atanassova *et al.* (2008) | 62 | 31.5 | 27.6 |  | 16.7 |  | 165 |  |
| Ichinose *et al*. (2018) | 71.5 | 10.4 | 23.6 |  | 38.4 | 74.4 |  |  |

DM = Diabetes Mellitus; HT = hypertension; SBP = systolic blood pressure

^a^Cell with double numbers means number for stroke, non-stroke sample. Cell with a single number means overall number.

**Table 3: Participants and study design.**

| **Author (Year)** | **Outcome** | **Specific study population** | **Study Design** | **Study Type** | **Mean Follow up time (year)** | **Number of subjects** | **Number of strokes** | **Type of dataset** |
| --- | --- | --- | --- | --- | --- | --- | --- | --- |
| Hung^30^ *et al*. (2018)^* †^ | Ischemic Stroke | Young | Retrospective Cohort | Derivation | 5 | 288582 | 215 | Multicenter |
| Chen-Ying^28^ *et al.* (2017)^*^ | Not specified or Both | General / Unspecified | Retrospective Cohort | Derivation | 5 | 798611 | 4944 | Multicenter |
| Kyriacou^53^ *et al.* (2015)^*^ | Ischemic Stroke | Carotid stenosis | Cohort | Derivation | > 3 | 1121 | 108 | Multicenter |
| Kastorini^35^ *et al.* (2013)^*^ | Ischemic Stroke | General / Unspecified | Nested Case-Control | Derivation |  | 250 | 250 | Multicenter |
| Hung^29^ *et al.*  (2019)^*†^ | Ischemic Stroke | General / Unspecified | Retrospective Cohort | Derivation | 3 | 840487 | 2544 | Multicenter |
| Chan^54^ *et al.* (2019)^*†^ | Ischemic Stroke | Recurrent stroke | Cohort | Derivation | 1 | 451 | 40 | Single center |
| Li^50^ *et al*. (2016)^*^ | Ischemic Stroke | AF | Retrospective Cohort | Derivation | 2 | 1864 | 193 | Multicenter |
| Orfanoudaki^39^ *et al*. (2020)^*^ | Not specified or Both | General / Unspecified | Retrospective Cohort | Derivation and External Validation | 10 | 18,793 | 1,013 | Multicenter |
| Ayyagari^55^ *et al*. (2014)^*^ | Ischemic Stroke | HT | Retrospective Cohort | Derivation | 5, 2, 3.7 | 36,117 | 4272 | Multicenter |
| Nwosu^24^ *et al.* (2019)^*†^ | Not specified or Both | General / Unspecified | Retrospective Cohort | Derivation |  | 29072 | 548 | Public dataset |
| Ray^27^ *et al*.(2020)^*†^ | Not specified or Both | General / Unspecified | Retrospective Cohort | Derivation |  | 29072 | 548 | Public dataset |
| Singh^38^ *et al*.  (2020)^*^ | Not specified or Both | Elderly | Nested Case-Control | Derivation |  | 1824 | 212 | Multicenter |
| Ahmed^21^ *et al*. (2019)^*†^ | Not specified or Both | General / Unspecified | Retrospective Cohort | Derivation |  | 43400 | 783 | Public dataset |
| Rado^26^ *et al*. (2019)^*†^ | Not specified or Both | General / Unspecified | Retrospective Cohort | Derivation |  | 43400 | 783 | Public dataset |
| Liu^23^ *et al*. (2019)^*†^ | Not specified or Both | General / Unspecified | Retrospective Cohort | Derivation |  | 43400 | 783 | Public dataset |
| Wu^56^ *et al.* (2020)^*^ | Not specified or Both | Elderly | Cohort | Derivation |  | 1131 | 56 | Multicenter |
| Zhang^40^ *et al*. (2019) | Ischemic Stroke | General / Unspecified | Retrospective Cohort | Derivation | 5 | 792 | 398 | Single center |
| Teoh^57^ (2018) | Not specified or Both | General / Unspecified | Nested Case-Control | Derivation | 1 | 8175 | 2725 | Single center |
| Yonglai^58^ *et al*. (2017) | Not specified or Both | General / Unspecified | Case-Control | Derivation |  | 792 | 398 | Single center |
| Zhang^59^ *et al.* (2019) | Ischemic Stroke | General / Unspecified | Case-Control | Derivation | 10 | 792 | 398 | Single center |
| Colak^60^ *et al*. (2015) | Not specified or Both | General / Unspecified | Case-Control | Derivation |  | 297 | 130 | Single center |
| Peng^25^ *et al*. (2020) | Not specified or Both | General / Unspecified | Retrospective Cohort | Derivation |  | 43400 | 783 | Public dataset |
| Li^61^ *et al.* (2017) | Ischemic Stroke | AF | Retrospective Cohort | Derivation | 1 | 6610 | 143 | Multicenter |
| Rosado^36^ *et al*. (2019) | Not specified or Both | General / Unspecified | Retrospective Cohort | Derivation |  | 1500 | 40 | Single center |
| Jeena^41^ *et al*. (2019) | Ischemic Stroke | General / Unspecified | Case-Control | Derivation |  | 820 | 492 | Single center |
| Zhang^62^ *et al*. (2018) | Not specified or Both | General / Unspecified | Case-Control | Derivation |  | 792 | 398 | Single center |
| Albu^63^ *et al*. (2019) | Not specified or Both | General / Unspecified | Retrospective | Derivation |  | 101 | NA | Single center |
| Songram^42^ *et al*. (2019) | Not specified or Both | Elderly | Case-Control | Derivation |  | 1000 | 500 | Single center |
| Sarihan^64^ *et al*. (2017) | Not specified or Both | General / Unspecified | Case-Control | Derivation |  | 208 | 104 | Single center |
| Kansadub^65^ *et al.* (2016)^†^ | Not specified or Both | General / Unspecified | Case-Control | Derivation |  | 750 | 250 | Single center |
| Clifford^32^ *et al*. (2019) | Not specified or Both | Certain country / Races | Retrospective Cohort | Derivation | 6 | 27744 | 460 | Multicenter |
| Arslan^66^ *et al*. (2016) | Ischemic Stroke | General / Unspecified | Case-Control | Derivation |  | 192 | 80 | Single center |
| Choi^22^ *et al*. (2020) | Not specified or Both | General / Unspecified | Retrospective Cohort | Derivation |  | 43400 | 783 | Public dataset |
| Qin^34^ *et al*. (2019) | Not specified or Both | Recurrent stroke | Retrospective Cohort | Derivation |  | 22538 |  | Multicenter |
| Peñafiel^67^ *et al.* (2018) | Not specified or Both | General / Unspecified | Retrospective Cohort | Derivation |  |  |  | Single center |
| Chantamit-O-Pas^68^ *et al*. (2017) | Not specified or Both | General / Unspecified | Retrospective Cohort | Derivation |  | 899 | 469 | Public dataset |
| Atanassova^31^ *et al.* (2008) | Ischemic Stroke | Recurrent stroke | Cohort | Derivation | 1 | 54 | 8 | Multicenter |
| Ichinose^33^ *et al*. (2018) | Ischemic Stroke | Carotid stenosis | Others | Derivation |  | 86 | 36 | Single center |
| Singh^37^ *et al*. (2017)^†^ | Not specified or Both | Elderly | Retrospective Cohort | Derivation |  | 1824 | 212 | Multicenter |
| Hu^69^ *et al*. (2020) | Not specified or Both | General / Unspecified | Retrospective Cohort | Derivation |  | 26 697 | 830 | Multicenter |

* A study in Low-RoB group

† Balancing dataset with up-sampling or down-sampling technique

**Table 4: Assessment RoB of each study by PROBAST domains**

| **ID** | **Participants RoB** | **Participants App.*** | **Predictors RoB** | **Predictors App.** | **Outcome RoB** | **Outcome App.** | **Analysis RoB** | **Overall judgment RoB** | **Overall judgment App.** |
| --- | --- | --- | --- | --- | --- | --- | --- | --- | --- |
| **1** | **Low** | **Low** | **Low** | **Low** | **Low** | **Low** | **Low** | **Low** | **Low** |
| **2** | **Low** | **Low** | **Low** | **Low** | **Low** | **Low** | **Low** | **Low** | **Low** |
| **3** | **Low** | **Low** | **Low** | **Low** | **Low** | **Low** | **Low** | **Low** | **Low** |
| **4** | **Low** | **Low** | **Low** | **Low** | **Low** | **Low** | **Low** | **Low** | **Low** |
| **5** | **Low** | **Low** | **Low** | **Low** | **Low** | **Low** | **Low** | **Low** | **Low** |
| **6** | **Low** | **Low** | **Low** | **Low** | **Low** | **Low** | **Low** | **Low** | **Low** |
| **7** | **Low** | **Low** | **Low** | **Low** | **Low** | **Low** | **Low** | **Low** | **Low** |
| **8** | **Low** | **Low** | **Low** | **Low** | **Low** | **Low** | **Low** | **Low** | **Low** |
| **9** | **Low** | **Low** | **Low** | **Low** | **Low** | **Low** | **Low** | **Low** | **Low** |
| **10** | **Unclear** | **Low** | **Low** | **Low** | **Low** | **Low** | **Unclear** | **Unclear** | **Low** |
| **11** | **Unclear** | **Low** | **Low** | **Low** | **Low** | **Low** | **Low** | **Unclear** | **Low** |
| **12** | **Low** | **Low** | **Low** | **Low** | **Unclear** | **Low** | **Unclear** | **Unclear** | **Low** |
| **13** | **Unclear** | **Low** | **Low** | **Low** | **Low** | **Low** | **Unclear** | **Unclear** | **Low** |
| **14** | **Unclear** | **Low** | **Low** | **Low** | **Low** | **Low** | **Unclear** | **Unclear** | **Low** |
| **15** | **Unclear** | **Low** | **Low** | **Low** | **Low** | **Low** | **Low** | **Unclear** | **Low** |
| **16** | **Low** | **Low** | **Low** | **Low** | **Low** | **Low** | **Unclear** | **Unclear** | **Low** |
| **17** | **High** | **Low** | **Low** | **Low** | **Low** | **Low** | **High** | **High** | **Low** |
| **18** | **Low** | **Low** | **Low** | **Low** | **Low** | **High** | **Low** | **Low** | **High** |
| **19** | **High** | **High** | **Low** | **Low** | **Low** | **Low** | **Unclear** | **High** | **Low** |
| **20** | **High** | **Low** | **Low** | **Low** | **Low** | **Low** | **Low** | **High** | **Low** |
| **21** | **High** | **Low** | **Low** | **Low** | **Low** | **Low** | **Low** | **High** | **Low** |
| **22** | **Low** | **Low** | **Low** | **Low** | **Low** | **Low** | **High** | **High** | **Low** |
| **23** | **Low** | **Low** | **Low** | **Low** | **Low** | **Unclear** | **Unclear** | **Unclear** | **Unclear** |
| **24** | **High** | **Low** | **Low** | **Low** | **Low** | **Low** | **High** | **High** | **Low** |
| **25** | **High** | **Low** | **Low** | **Low** | **Low** | **Low** | **Low** | **High** | **Low** |
| **26** | **High** | **Low** | **Low** | **Low** | **Low** | **Low** | **Low** | **High** | **Low** |
| **27** | **High** | **Unclear** | **Unclear** | **Unclear** | **Low** | **Unclear** | **High** | **High** | **Unclear** |
| **28** | **High** | **Low** | **Low** | **Low** | **Low** | **Unclear** | **Unclear** | **High** | **Unclear** |
| **29** | **High** | **Unclear** | **Low** | **Low** | **Low** | **Low** | **Unclear** | **High** | **Unclear** |
| **30** | **Unclear** | **High** | **Low** | **Low** | **Low** | **Low** | **Low** | **Unclear** | **High** |
| **31** | **Low** | **Low** | **Low** | **High** | **Low** | **Low** | **Unclear** | **Unclear** | **High** |
| **32** | **High** | **Low** | **Low** | **Unclear** | **Low** | **Low** | **Unclear** | **High** | **Unclear** |
| **33** | **Unclear** | **Unclear** | **Low** | **Unclear** | **Unclear** | **Unclear** | **High** | **High** | **High** |
| **34** | **High** | **Low** | **Low** | **Low** | **Unclear** | **High** | **High** | **High** | **High** |
| **35** | **Low** | **Low** | **Low** | **Low** | **High** | **High** | **Unclear** | **High** | **High** |
| **36** | **Unclear** | **High** | **Low** | **Low** | **Unclear** | **High** | **High** | **High** | **High** |
| **37** | **Low** | **Low** | **Low** | **Low** | **Low** | **Low** | **High** | **High** | **High** |
| **38** | **Low** | **Low** | **High** | **Low** | **High** | **High** | **High** | **High** | **High** |
| **39** | **High** | **High** | **High** | **High** | **High** | **High** | **High** | **High** | **High** |
| **40** | **Low** | **Low** | **Unclear** | **Unclear** | **Low** | **Low** | **Unclear** | **High** | **High** |

* App.: Applicability

**Table 5: Evaluation metrics.**

| **Author (Year)** | **Feature selection** | **Group of models** | **Type of Model (for ML)** | **AUC** | **Sensitivity / Recall** | **Specificity** | **Precision** | **F1-score** | **Accuracy** |
| --- | --- | --- | --- | --- | --- | --- | --- | --- | --- |
| Hung *et al*. (2018) | Pearson correlation | Neural network | Neural network | 0.8 |  |  |  |  |  |
| Chen-Ying *et al*. (2017) | Information gain (Entropy / Gini) | Boosting | Gradient Boosting Decision Tree (GBDT) | 0.918 | 0.856 | 0.865 |  |  | 0.868 |
|  |  | Neural Network | Deep Neural network | 0.915 | 0.845 | 0.871 |  |  | 0.873 |
|  |  | Regression-based | Logistic regression |  | 0.82 | 0.864 |  |  | 0.866 |
|  |  | SVM | SVM |  | 0.813 | 0.837 |  |  | 0.839 |
| Kyriacou *et al.* (2015) |  | SVM | SVM | 0.81 (0.80-0.82) | 0.88 (0.82-0.94) | 0.72 (0.66-0.78) |  |  | 0.78 (0.71-0.85) |
| Kastorini *et al.* (2013) | PCA | Naive Bayes | Naive Bayes | 0.77 |  |  |  |  |  |
|  |  | Regression-based | Multiple Logistic Regression | 0.78 |  |  |  |  |  |
|  |  | Decision Tree | C4.5 | 0.617 |  |  |  |  |  |
|  |  | Rule Induction | RIPPER | 0.665 |  |  |  |  |  |
|  |  | SVM | SVM | 0.684 |  |  |  |  |  |
|  |  | Neural network | Multilayer perceptron | 0.729 |  |  |  |  |  |
| Hung *et al*. (2019) | univariate Pearson correlation | Neural network | Neural network | 0.925 | 0.92 | 0.8 |  |  |  |
| Chan *et al*. (2019) |  | Neural network | Neural network | 0.77 (0.68-0.84) | 0.75 (0.63-0.83) | 0.75 (0.63-0.83) |  |  | 0.75 (0.69-0.77) |
|  |  | SVM | SVM | 0.77 (0.68-0.84) | 0.625 (0.5-0.625) | 0.75 (0.50-0.75) |  |  | 0.625 (0.563-0.688) |
|  |  | Naive Bayes | Naive Bayes | 0.77 (0.68-0.84) | 0.625 (0.5-0.625) | 0.75 (0.63-0.83) |  |  | 0.625 (0.563-0.688) |
| Li *et al*. (2016) | Comparing between various method (Chi-square, correlation-based feature subset selection method (CFS), wrapper for AUC, Information gain, Lasso) | Regression-based | Logistic regression | 0.759 |  |  |  |  |  |
|  |  | Regression-based | Cox proportional hazards regression | 0.735 |  |  |  |  |  |
|  |  | Naive Bayes | Naive Bayes | 0.745 |  |  |  |  |  |
|  |  | Decision Tree | CART | 0.667 |  |  |  |  |  |
|  |  | Random Forest | Random Forest | 0.757 |  |  |  |  |  |
| Orfanoudaki *et al*. (2020) | Information gain (Entropy / Gini) | Decision Tree | Optimal Classification Trees | 0.8743 (0.8569–0.9014) | 0.9142 | 0.7238 | 0.9408 |  |  |
|  |  | Regression based | Logistic regression | 0.8065 (0.772–0.8351) | 0.8933 | 0.7102 | 0.9701 |  |  |
|  |  | Decision Tree | CART | 0.7981 (0.7676–0.8287) | 0.8802 | 0.7099 | 0.9736 |  |  |
|  |  | Random Forest | Random Forest | 0.8829 (0.8578–0.9081) | 0.9175 | 0.7161 | 0.9605 |  |  |
|  |  | Boosting | XGBoost | 0.8846 (0.8643–0.9048) | 0.9167 | 0.7354 | 0.9412 |  |  |
| Ayyagari *et al*. (2014) | LASSO |  | Regression-based | 0.729 |  |  |  |  |  |
| Nwosu *et al.* (2019) | PCA | Neural network | Neural network |  |  |  |  |  | 0.7502 |
|  |  | Random Forest | Random Forest |  |  |  |  |  | 0.7453 |
|  |  | Decision Tree | Decision Tree |  |  |  |  |  | 0.7431 |
| Ray *et al*. (2020) | Chi-square | Random Forest | Decision jungle | 0.77 | 0.058 |  | 0.073 | 0.065 | 0.967 |
|  |  | Boosting | Boosted Decision Tree | 0.78 | 0.025 |  | 0.09 | 0.37 | 0.976 |
| Singh *et al.* (2020) | Information gain (Entropy / Gini), PCA | SVM | SVM |  |  |  |  |  | 0.94 |
|  |  | Neural network | Neural network |  |  |  |  |  | 0.786 |
|  |  | Neural network | PCA + Neural network |  |  |  |  |  | 0.929 |
|  |  | Stacking | Decision Tree + Neural network |  |  |  |  |  | 0.929 |
|  |  | Stacking | Decision Tree + PCA + Neural network |  | 1 | 0.944 |  |  | 0.952 |
| Ahmed *et al.* (2019) |  | Regression-based | Logistic regression |  | 0.82 |  | 0.75 | 0.79 | 0.9 |
|  |  | Random Forest | Random Forest |  | 0.96 |  | 0.87 | 0.91 | 0.77 |
|  |  | Decision Tree | Decision Tree |  | 0.84 |  | 0.77 | 0.81 | 0.79 |
|  |  | SVM | SVM |  | 0.83 |  | 0.75 | 0.79 | 0.77 |
| Rado *et al.* (2019) |  | KNN | KNN |  |  |  | 0.846 | 0.846 | 0.8458 |
|  |  | SVM | SVM |  |  |  | 0.777 | 0.772 | 0.7729 |
|  |  | Decision Tree | Decision Tree |  |  |  | 0.861 | 0.861 | 0.861 |
|  |  | Random Forest | Random Forest |  |  |  | 0.867 | 0.866 | 0.8663 |
|  |  | Boosting | AdaBoost |  |  |  | 0.825 | 0.824 | 0.8243 |
|  |  | Stacking | Stacking |  |  |  | 0.876 | 0.876 | 0.8758 |
| Liu *et al.* (2019) | PCA | Neural network | Automated Hyperparameter Optimization (AutoHPO) + Neural network |  | 0.674 | 0.326 |  |  | 0.716 |
|  |  | Neural network | Neural network |  | 0.677 | 0.323 |  |  | 0.651 |
|  |  | Random Forest | Bagging |  | 0.876 | 0.124 |  |  | 0.738 |
|  |  | Random Forest | Random Forest |  | 0.893 | 0.107 |  |  | 0.728 |
|  |  | Boosting | XGBoost |  | 0.887 | 0.113 |  |  | 0.741 |
|  |  | Boosting | AdaBoost |  | 0.888 | 0.112 |  |  | 0.733 |
| Wu *et al.* (2020) |  | Regression based | Logistic regression | 0.72 (0.71-0.73) | 0.75 (0.72-0.78) | 0.69 (0.67-0.71) |  |  | 0.7 (0.68-0.72) |
|  |  | Random Forest | Random Forest | 0.71 (0.70-0.72) | 0.62 (0.58 -0.66) | 0.79 (0.77-0.81) |  |  | 0.78 (0.77-0.79) |
|  |  | SVM | SVM | 0.71 (0.70-0.72) | 0.7 (0.66-0.74) | 0.72 (0.68-0.76) |  |  | 0.72 (0.68-0.76) |
| Zhang *et al.* (2019) | Information gain (Entropy / Gini) | SVM | SVM |  | 0.827 | 0.804 |  |  | 0.815 |
|  |  | Naive Bayes | Naive Bayes |  | 0.802 | 0.5 |  |  | 0.657 |
|  |  | Others | Classification based on association |  | 0.766 | 0.761 |  |  | 0.764 |
|  |  | Neural network | Neural network |  | 0.704 | 0.558 |  |  | 0.632 |
|  |  | Decision Tree | CART |  | 0.704 | 0.718 |  |  | 0.71 |
|  |  | Decision Tree | C4.5 |  | 0.676 | 0.766 |  |  | 0.721 |
| Teoh (2018) |  | Neural network | Neural network | 0.67 |  |  |  |  |  |
| Yonglai *et al*. (2017) | Standard Deviation Ranking and voting | SVM | Glowworm swarm optimization (GSO) with SVM |  |  |  |  |  | 0.7323 |
|  | wrapper | SVM | SVM |  |  |  |  |  | 0.6312 |
|  |  | Neural network | Neural network |  |  |  |  |  | 0.6863 |
| Zhang *et al.* (2019) | Relief algorithm | SVM | SVM |  |  |  |  |  | 0.77 |
| Colak *et al*. (2015) | Cramer's V test | SVM | SVM | 0.91 |  |  |  |  | 0.8462 |
|  |  | Neural network | Neural network | 0.928 |  |  |  |  | 0.859 |
| Peng *et al*. (2020) |  | Neural network | Neural network |  |  |  |  |  | 0.98 |
| Li *et al*. (2017) | Boots-wrapper | Regression-based | Regression-based | 0.714 |  |  |  |  |  |
| Rosado *et al*. (2019) | PCA | SVM | SVM |  | 0.81 | 0.99 | 0.75 | 0.78 | 0.99 |
| Jeena *et al*. (2019) | Chi-square | Regression-based | Regression-based | 0.97 | 0.96 |  | 0.99 | 0.97 | 0.97 |
| Zhang *et al*. (2018) |  | SVM | SVM | 0.8948 |  |  |  |  | 0.8258 |
| Albu *et al*. (2019) |  | Neural network | Neural network |  |  |  |  |  | 0.89 |
| Songram *et al*. (2019) | Chi-square | KNN | KNN |  | 0.3716 |  | 0.5734 | 0.4476 | 0.547 |
| Songram *et al.* (2019) | Chi-square | Naive Bayes | Naive Bayes |  | 0.6765 |  | 0.5707 | 0.6164 | 0.585 |
| Songram *et al.* (2019) | Chi-square | SVM | SVM |  | 0.5372 |  | 0.6713 | 0.585 | 0.622 |
| Songram *et al*. (2019) | Chi-square | Decision Tree | Decision Tree |  | 0.5001 |  | 0.6012 | 0.5379 | 0.575 |
| Sarihan *et al*. (2017) |  | SVM | SVM | 0.97 |  |  |  |  | 0.87 |
| Kansadub *et al*. (2016) | Specialist | Naive Bayes | Naive Bayes | 0.769 |  |  |  |  | 0.72 |
| Kansadub *et al*. (2016) | Specialist | Decision Tree | Decision Tree | 0.725 |  |  |  |  | 0.75 |
| Kansadub *et al.* (2016) | Specialist | Neural network | Neural network | 0.75 |  |  |  |  | 0.74 |
| Clifford *et al*. (2019) | Univariate with standard Backward-Forward elimination | Regression-based | Logistic regression |  |  |  |  |  |  |
|  |  | Naive Bayes | Naive Bayes |  |  |  |  |  |  |
|  |  | Neural Network | Neural network |  |  |  |  |  |  |
|  |  | Random Forest | Random Forest |  |  |  |  | 0.91 |  |
| Arslan *et al*. (2016) |  | SVM | SVM | 0.9783 (0.9569–0.9997) | 0.9747 (0.9115–0.9969) | 0.9820 (0.9364–0.9978) |  |  | 0.9789 (0.9740–0.9942) |
|  |  | Boosting | Stochastic Gradient Boosting | 0.9757 (0.9543–0.9970) | 0.9512 (0.8797–0.9865) | 0.9907 (0.9494–0.9997) |  |  | 0.9737 (0.9397–0.9914) |
|  |  | Regression based | Penalized Logistic Regression | 0.8953 (0.8510–0.9396) | 0.8554 (0.7610–0.9230) | 0.9252 (0.8579–0.9671) |  |  | 0.8947 (0.8421–0.9345) |
| Choi *et al*. (2020) |  | Decision Tree | Decision Tree |  | 0.04 |  | 0.16 |  |  |
| Qin *et al.* (2019) | Univariate with standard Backward-Forward elimination | SVM | SVM |  | 0.96 |  | 0.93 |  |  |
|  |  | Neural network | LSTM |  | 0.97 |  | 0.95 |  |  |
| Peñafiel *et al*. (2018) |  | Naive Bayes | Naive Bayes |  |  |  |  |  | 0.6 |
|  |  | Neural network | Neural network |  |  |  |  |  | 0.6 |
|  |  | Others | Dempster-Shafer Theory |  |  |  |  |  | 0.6 |
|  |  | Others | Quadratic Discriminant Analysis |  |  |  |  |  | 0.6 |
| Chantamit-O-Pas *et al*. (2017)^*^ | From AHA guideline | Naive Bayes | Naive Bayes |  |  |  |  |  |  |
|  |  | SVM | SVM |  |  |  |  |  |  |
|  |  | Neural network | Neural network |  |  |  |  |  |  |
| Atanassova *et al.* (2008) | Univariate with standard Backward-Forward elimination | Neural network | Neural network | 0.79 (0.6-0.99) |  |  |  |  | 0.8 |
| Ichinose *et al.* (2018) | Univariate with standard Backward-Forward elimination | Neural network | Neural network | 0.768 | 0.5 | 0.82 |  |  | 0.686 |
| Singh *et al.* (2017) | Decision tree + PCA for feature selection | Neural network | Neural network |  |  |  |  |  | 0.977 |
| Hu *et al.* (2020) ^†^ |  |  | Regression-based, Naive Bayes, Random Forest, Boosting, Bayesian additive regression trees (BART) |  |  |  |  |  |  |

CART = Classification and regression trees; SVM = Support vector machine; PCA = Principal component analysis; KNN = k-Nearest neighbors; LSTM = Long short-term memory; RIPPER = Repeated Incremental Pruning to Produce Error Reduction

* Report mean-square error

† Aim to compare top-five features between models

**Table 6: Important features in the studies**

| **Features** | **Frequency** |
| --- | --- |
| Age | 12 |
| Gender | 9 |
| Smoking | 7 |
| Hypertension | 7 |
| Heart disease (including Myocardial Infarction) | 7 |
| Body Mass Index | 6 |
| Diabetes Mellitus | 5 |
| Blood Glucose | 5 |
| Work-type | 5 |
| Dyslipidemia | 4 |
| Marital status | 4 |
| Thromboembolism | 2 |
| Systolic Blood Pressure | 2 |
| High-density lipoprotein cholesterol (HDL) | 2 |
| Serum creatinine | 2 |
| Education | 2 |
